# Paediatric Palliative Care following Hospital discharge: Prevalence and factors associated with non-continuity of palliative care for children with cancer in Busoga sub-region-eastern Uganda; A mixed methods study

**DOI:** 10.1101/2025.01.07.25320167

**Authors:** Miriam Ajambo, Joseph Rujumba, Savio Mwaka, Joseph Gavin Nyanzi, Damalie Nalwanga, Joyce Balagadde

## Abstract

**Background:** Palliative care (PC) is crucial for children with cancer to alleviate suffering and enhance quality of life. However, continuity of pediatric palliative care (PPC) can be disrupted by factors such as lack of knowledge, stigma, bureaucratic hurdles, inadequate referral systems, and staffing shortages. There is limited data on the prevalence and factors associated with non-continuity of PPC in Uganda. This study explores the prevalence and factors contributing to non-continuity of PC among children with cancer in Uganda with Busoga Region in Eastern Uganda as a case study.

**Methodology:** This cross-sectional mixed-methods study was conducted at two specialized tertiary facilities in Uganda managing pediatric cancer. Data were extracted from online databases for 307 children treated from 2019 to 2023, of whom 80 were alive during the study. Caregivers of 77 children participated in interviews, and nine key informants from the two facilities and the only regional facility providing PC upon downward referral were also interviewed. Descriptive statistics summarized data as proportions or percentages, and bivariate analysis used crude odds ratios to identify significant associations. Key informant interviews were transcribed and analyzed thematically using the socio-ecological model.

**Results:** The prevalence of non-continuity of PC was 96.1% (95% CI: 88.4-98.0). Barriers identified included: individual-level gaps in caregiver knowledge; relationship-level issues such as inappropriate cultural beliefs; health system-level challenges like limited human resources, inadequate training and funding, poor coordination and referral pathways, and service access issues; and policy-level concerns, including the lack of a national palliative care policy.

**Conclusion:** The high prevalence of non-continuity of PC for children with cancer in Busoga highlights significant deficiencies in integrating palliative care into pediatric oncology services in Uganda. Addressing these challenges requires urgent government action to enhance palliative care funding and resources.

## Introduction

Palliative care (PC) employs a holistic approach to improve the quality of life for patients suffering from serious illnesses like cancer, as well as their families, by addressing the psychosocial, spiritual, and physical challenges associated with the illness. Suffering is prevented or relieved through early diagnosis and appropriate management of pain and other distressing symptoms [1]. The World Health Organization (WHO) describes paediatric palliative care (PPC) as the total care of a child’s body, mind, and spirit [2]. PPC includes pain management, counselling, specialized care, and psychosocial and spiritual support [3].

Cancer is an increasing public health burden in Sub-Saharan Africa [3]. According to the WHO Global Initiative on Childhood Cancers 2018, approximately 400,000 children are diagnosed with cancer annually, with 80% of these cases occurring in low– and middle-income countries (LMICs). Following diagnosis, all these children require palliative care, making the integration of PPC into standard paediatric cancer care critically important [4, 5]. However, one major challenge in LMICs is the difficulty in accessing PC, which hampers the continuity of services upon patient discharge [4]. Palliative care is considered best practice and is increasingly implemented earlier in the trajectory of cancer [6, 7]. To ensure adequate symptom management, prevent patient suffering, and establish a relationship of trust, continuity of PPC after discharge from the hospital is key. There should not be a gap in continuity for more than 14 days following discharge [1]. Access to PPC by children in LMICs is affected by low levels of awareness among health workers about palliative care and an inadequate number of palliative care service providers [8]. In some African countries, such as Nigeria and Morocco, failure to refer and late referral are the main service factors known to influence symptom control and cause distress among caregivers [9].

In Uganda, government support for PC has grown over the years, although a comprehensive strategy and guidelines are not yet in place. Currently, there are 13 PC centers that support the two tertiary facilities—Uganda Cancer Institute (UCI) and Mulago National Referral Hospital Paediatric Haematology-Oncology Unit (MNRH-PHOU)—in providing continuity of PPC. The limited information on factors associated with non-continuity of PPC in Uganda prompted this study.

There is an increasing number of children with cancer in Uganda, with 1,000 cases diagnosed annually, 750 of which are managed at UCI, and about 15.6% of these cases are from the Busoga sub-region in Eastern Uganda [10]. There is hardly any data on the non-continuity of PPC after discharge. This study aimed to establish the prevalence and factors associated with the non-continuity of PPC in Uganda.

## Materials and methods

### Study design

This study employed a cross-sectional mixed methods sequential explanatory analytical design. It was conducted at two tertiary facilities providing pediatric palliative care (PPC) and one palliative care institution where downward referrals from the two facilities are directed. A search of the hospital electronic databases at the two tertiary facilities was conducted for children who presented from 2019 to 2023. Caregivers of these children and health workers providing PPC at the facilities were interviewed. Quantitative data collection and analysis preceded the qualitative. Caregivers were contacted by phone to ascertain whether the child was continuing PC and were then invited to respond to a questionnaire. Key informant interviews were conducted, recorded, and transcripts were generated.

Ethical approval was obtained from the School of Medicine Research and Ethics Committee (SOMREC). Patients’ records were handled with confidentiality and privacy, and informed consent was obtained from all participants.

### Study setting and participants

The population of interest included children with cancer from the Busoga sub-region in Eastern Uganda who presented from 2019 to 2023, and health workers providing PPC at the two tertiary facilities and the palliative care institution in the sub-region. There are an estimated 2,392,450 children in this region, representing 10.3% of the population of all children in Uganda, translating to approximately 332 cases of children with cancer [11]

Inclusion Criteria; Caregivers of children who had been discharged from the two tertiary facilities within the study period and were documented residents of the Busoga sub-region, as well as palliative care service providers at the three facilities who consented to participate in the study.

Exclusion Criteria; Caregivers of children who died after discharge, those without telephone contacts, or those who were unreachable. All PC providers not involved in PPC and PC providers at the three facilities who had not worked at the facility for at least two years.

### Data collection procedures

Quantitative Data Collection; Data was collected using a semi-structured questionnaire through Kobo Collect, an online data collection kit. The questionnaire explored patient demographics, and patient-level, health system, and policy factors associated with non-continuity of PPC.

Qualitative Data Collection; Key informants were interviewed at their respective places of work. These interviews were recorded and transcribed verbatim. The key informant interview guide explored health system, patient-level, and policy-related factors.

Data Quality Control; Data collection tools were pretested. Online data collection facilitated near real-time record review. Daily data reviews were conducted and corrective actions were taken. The team that transcribed the interviews was different from the team that conducted them to avoid interviewer bias and ensure consistency between the transcripts and recordings.

### Data analysis

Quantitative data analysis was performed using STATA 15 [12]. Descriptive statistics for categorical data were summarized as proportions or percentages and presented using tables. Crude odds ratios were used to establish bivariate relationships. The prevalence of non-continuity was calculated as a proportion, with the numerator being the number of children who did not continue PC and the denominator being the total number of children in the study.

Qualitative data analysis was conducted manually. Transcripts were read multiple times to identify codes from which a codebook was generated, detailing facilitators and barriers. These barriers and facilitators were analyzed using the socio-ecological model (SEM), exploring factors at the individual, relationship, health system, and policy levels [13].

## Results

A total of 307 children were discharged from the two tertiary facilities between 2019 and 2023. Of these, 153 (49.8%) had died, and 74 (24.1%) were unreachable. Among the 80 eligible participants, 77 consented and were enrolled.

**Figure 1.**
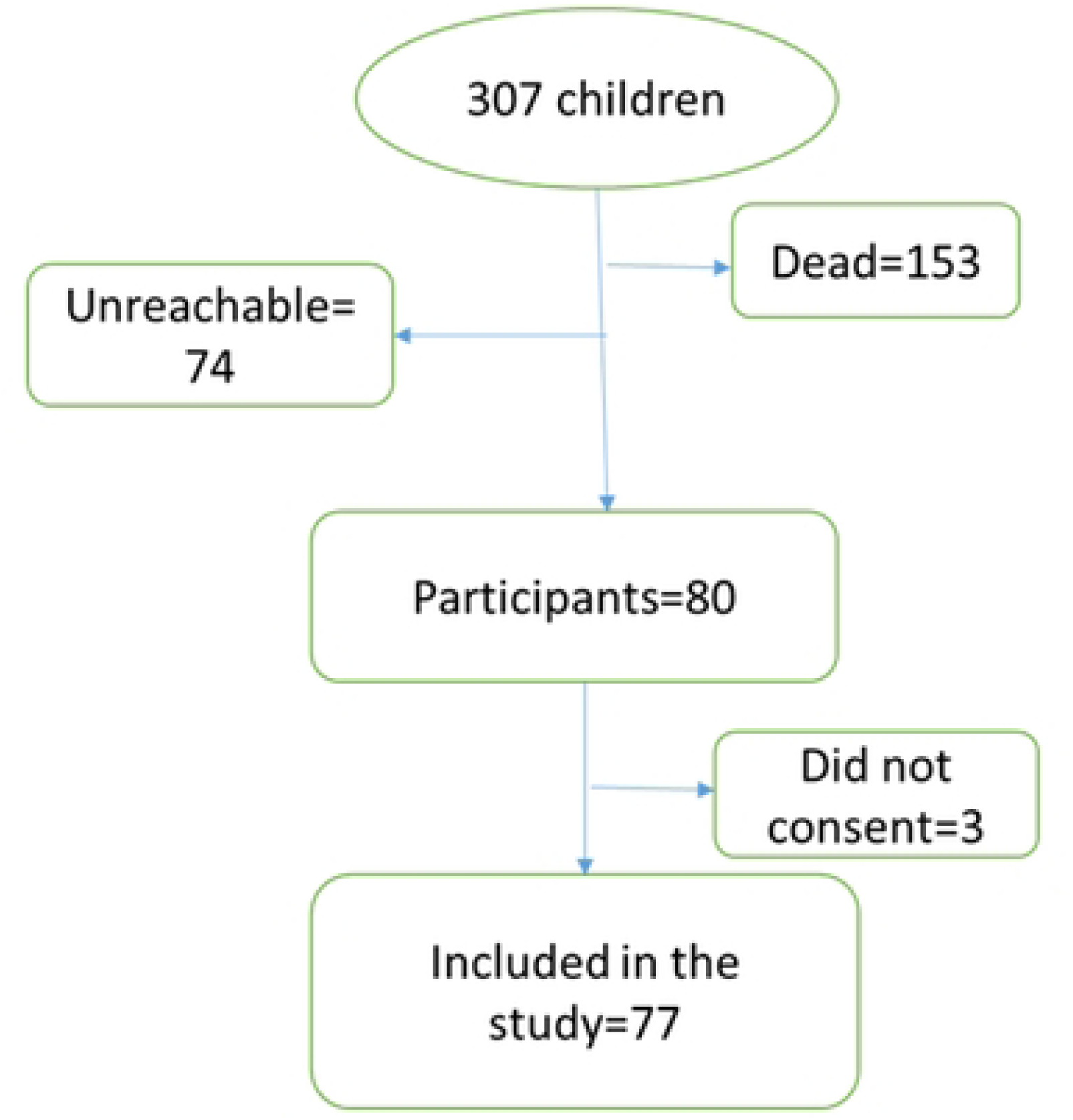
illustrates the enrollment flow.

### Prevalence of non-continuity of PC

Of the 77 caregivers enrolled, 74 (96.1%; CI: 88.4 –98.8) reported that their children did not continue to receive PC services.

Relationship between patient characteristics and non-continuity of PPC Seventy-four out of 77(96.1%) children did not continue to receive palliative care after discharge, 36/74(48.6%) of these were not in school. However, all three who continued to receive PC were also not in school. Of those that did not continue, 33/74(44.6%) had liquid tumours and of these, 20/33(60.6%) had non-Hodgkin’s lymphoma, predominantly Burkitts Lymphoma. None of the characteristics showed a statistically significant relationship. Table one shows the relationship between patient characteristics and non-continuity.

**Table 1:** Relationship between patient characteristics and non-continuity.

| **Variable** | **continued,**<br>**N=3(3.9%),**<br>**n(column%)** | **Didn't continue ,**<br>**N=74, n(column%)** | **COR(95% CI)** | **p value** |
| --- | --- | --- | --- | --- |
| **Age** |  |  |  | 0.800 |
| 0-4 years | 0(0.0) | 21(28.4) | 1.000 |  |
| 5-9 years | 1(33.3) | 21(28.4) | 0.429(0.024,7.632) | 0.564 |
| 10-14 years | 1(33.3) | 23(31.1) | 0.391(0.022,6.949) | 0.523 |
| 15-17 years | 1(33.3) | 9(12.2) | 1.000 |  |
| **Gender** |  |  |  | 0.801 |
| Female | 1(33.3) | 30(40.5) | 1.000 |  |
| Male | 2(66.7) | 44(62.9) | 1.364(0.118,15.722) | 0.804 |
| **Diagnosis** |  |  |  |  |
| **Solid tumours** |  |  |  |  |
| Bone tumour | 0(0.0) | 1(1.4) |  |  |
| Soft tissue sarcoma | 0(0.0) | 9(12.2) |  |  |
| Brain tumour | 0(0.0) | 5(6.8) |  |  |
| Wilm's tumour | 0(0.0) | 14(18.9) |  |  |
| **Liquid tumours** |  |  |  |  |
| Leukaemia | 0(0.0) | 13(17.6) |  |  |
| Non-Hodgkin's | 0(0.0) | 20(26.0) |  |  |
| Others | 3(100) | 12(16.2) |  |  |
| **Child's class** |  |  |  |  |
| Not in school | 3(100) | 36(48.6) |  |  |
| Pre-primary | 0(0.0) | 7(9.5) |  |  |
| Lower primary | 0(0.0) | 18(24.3) |  |  |
| Upper primary | 0(0.0) | 13(17.6) |  |  |
| **Duration of symptoms** |  |  |  | 0.362 |
| < 3 months | 0(0.0) | 29(39.2) | 1.000 |  |
| 3-<6 months | 2(66.7) | 13(17.6) | 3.077(0.253,37.483) | 0.378 |
| 6< 1 year | 0(0.0) | 12(16.2) | 1.000 |  |
| 1 year + | 1(33.3) | 20(27.0) | 1.000 |  |

Relationship Between Caregiver Characteristics and Non-Continuity of PPC Seventy out of 77 (90.9%) caregivers in the study were biological parents. Among these, 95.7% did not continue with PC. All children whose caregivers were not their biological parents did not continue with PC. None of the caregiver characteristics showed a statistically significant relationship. Table 2 presents the caregiver characteristics and their relationship with non-continuity of PC

**Table 2:** Relationship between caregiver characteristics and non-continuity.

| **Variable** | **continued,**<br>**N=3(3.9%),**<br>**n(column%)** | **Didn't**<br>**continue ,**<br>**N=74,**<br>**n(column%)** | **N=77(100%)** | **COR(95%CI)** | **p** |
| --- | --- | --- | --- | --- | --- |
| **Caretaker relationship** |  |  |  |  |  |
| Parent | 3(100.0) | 67(90.5) | 70(90.9) |  |  |
| Grandfather | 0(0.0) | 4(5.5) | 4(5.2) |  |  |
| Other relatives | 0(0.0) | 3(4.1) | 2(2.6) |  |  |
| **Caretaker marital status** |  |  |  |  |  |
| Married/cohabiting | 3(100) | 63(85.1) | 66(85.7) |  |  |
| Separated/divorced | 0(0.0) | 5(6.8) | 5(6.5) |  |  |
| Single | 0(0.0) | 4(5.4) | 4(5.2) |  |  |
| Widowed | 0(0.0) | 2(2.7) | 2(2.6) |  |  |
| **Education level of household head** |  |  |  |  | 0.792 |
| No formal education | 0(0.0) | 7(9.5) | 7(9.1) | 1.000 |  |
| Primary education | 2(66.7) | 32(43.2) | 34(44.2) | 1.375(0.117,16.111) | 0.800 |
| Secondary education | 1(33.3) | 22(29.7) | 23(29.9) | 1.000 |  |
| Tertiary education | 0(0) | 13(17.6) | 13(16.9) | 1.000 |  |
| **Caregiver occupation** |  |  |  |  | 0.414 |
| Business person | 1(33.3) | 8(10.8) | 9(11.7) | 1.000 |  |
| Civil servant | 0(0.0) | 6(8.1) | 6(7.8) | 1.000 |  |
| Others | 0(0.0) | 11(14.9) | 11(14.3) | 1.000 |  |
| Peasant | 2(66.7) | 49(66.2) | 51(66.2) | 0.383(0.026,4.034) | 0.328 |
| **Setting** |  |  |  |  |  |
| Rural | 3(100) | 42(56.8) | 45(58.4) |  |  |
| Urban | 0(0.0) | 32(43.2) | 32(41.6) |  |  |

### Relationship between health system factors and non-continuity of PPC

Those who did not receive PC information from Health workers had higher odds COR; 22.667(95% CI 1.785,287.820), p= 0.016 of non-continuity of PC. All three that continued had been given PC information by the healthcare workers.

All the 74 that did not continue PC were not given referral information for continuity of PC at discharge but all the three that continued received it. Two were referred to RHHJ and one, to Jinja RRH.

### Qualitative analysis results from key informant interviews

Nine KI were selected from the three study sites. Table 3 presents the demographic characteristics of the key informants

**Table 3:** Demographic characteristics of the Key informants.

|  | MNRH-PHOU<br>N=3(33.3%) | UCI<br>N=3(33.3%) | RHHJ<br>N=3(33.3%) | N=9(100%) |
| --- | --- | --- | --- | --- |
| **Sex** |  |  |  |  |
| Male | 1 | 1 | 0 | 2(22.2) |
| Female | 2 | 2 | 3 | 7(77.8) |
| **Years in service** |  |  |  |  |
| < 5 | 1 | 1 | 0 | 2(22.2) |
| 5-9 | 2 | 1 | 2 | 5(55.6) |
| 10-14 | 0 | 1 | 1 | 2(22.2) |
| **Designation** |  |  |  |  |
| Nursing officer | 1 | 1 | 0 | 2(22.2) |
| Social worker | 0 | 1 | 1 | 2(22.2) |
| PC clinical Officer | 0 | 0 | 2 | 2(22.2) |
| Medical officer | 1 | 1 | 0 | 2(22.2) |
| Paediatric haematologist oncologist | 1 | 0 | 0 | 1(11.1) |
| **Palliative care training** |  |  |  |  |
| Diploma | 2 | 1 | 3 | 6(66.7) |
| certificate | 1 | 1 | 0 | 2(22.2) |
| No training | 0 | 1 | 0 | 1(11.1) |

### Theme 1: Informants’ understanding of PC

#### Sub-theme; Good understanding of PC

Most of the key informants had good understanding of what palliative care is. They defined it as; an approach which gives quality of care to the patient with life-threatening illnesses, an art of routine medical care that should be given to all patients with either chronic conditions or life-threatening conditions. It entails looking at those social aspects of life, the cultural stressors and the support that one gives to the patient and the family from the time of diagnosis till death.

> *“It is an approach which gives quality of care to the patient with life-threatening illnesses; patients and their families, identifying their symptoms, and managing the pain is very important, then following them up through diagnosis to the end-of-life” **-KII 1, MNRH***

#### Sub-theme 2; Poor understanding of PC

One participant had an idea of what PC is, she could not define it comprehensively and only knew it as care at the end of one’s life.

> *“Ideally, I would take palliation as the care at the end of life for someone who we believe we are not going to do much in terms of curing them. We don’t start it at the start of treatment”**-KII 6, UCI***

#### Theme 2: Barriers and Facilitators to Continuity of PC for Children with Cancer in the Busoga Sub-Region

The study identified factors that positively influenced (facilitators) and negatively influenced (barriers) the continuity of PPC. These factors were analyzed and presented in line with the socio-ecological model (SEM) adopted from Pierce and Kealey, 2023 (49). The sub-themes and codes at the individual, relationship, health system, and societal levels regarding facilitators and barriers to the continuity of PPC are illustrated in the figure below.

### Individual-Level Factors Hindering Continuity of PC for Children with Cancer in the Busoga Sub-Region

#### Knowledge Gap on Palliative Care Among Caregivers

A knowledge gap was a key barrier to the continuity of PPC. Key informants noted that caregivers cannot adopt practices they are unaware of.

> *“Sometimes it is illiteracy, most patients’ attendants may not know that palliative care can help somebody to have a quality life or live long” **KII 2, MNRH***

#### Childhood Developmental Aspects

A few key informants reported that children’s expressions differ from those of adults, posing a challenge in the continuity of palliative care. One key informant stated that pain assessment and grading in children are difficult, which hinders the continuity of PPC.

> *“It is very hard to understand how children experience pain! Sometimes the child is crying and you think it’s pain yet the child needs to shower! So those elements make it very hard to know the level of pain”-**KII 4, UCI**.*

### Relationship Barriers Influencing Continuity of PC for Children in the Busoga Sub-Region

#### Social Challenges

A few key informants highlighted social challenges. One key informant noted that a cancer diagnosis can lead to parental separation or divorce, which itself hinders the continuity of PPC. Palliative care for a child requires full family support, and a fragmented family can face challenges such as interruptions in care-seeking and difficulties in raising finances to transport the child to receive the necessary services

> *“Sometimes, most patients have social problems-when a child is diagnosed with cancer, the man will divorce the mother saying they don’t give birth to cancer patients. When the family is shattered it affects the continuity” –**KII 1, MNRH***

### Inappropriate Cultural Beliefs

Some key informants cited that certain cultural beliefs among caregivers hinder the continuity of PPC. It was noted that some caregivers attribute the cause of cancer to witchcraft, leading them to seek out witch doctors instead of continuing with medical services. Key informants also mentioned that some caregivers believe cancer could result from infidelity.

> *“Beliefs; some don’t believe in palliative care, so after you have given them advice, you link them up but when they go, they do something else. Most of them think it [cause of cancer] is witchcraft and when they are discharged from here, they go and look for other ways-they go to witch doctors. I got a family that had separated because the child had cancer and the mother was accusing the dad that this child got cancer because he cheated! During bereavement, it took us time for almost a year and half to ensure that these people were back together and I was happy for the success”-**KII 3, MNRH***

### Health System-Related Barriers to Continuity of PC

#### Limited Human Resources for Pediatric Palliative Care

Most key informants described how working alone in the palliative care unit is overwhelming and hinders the continuity of services. They also reported having many other responsibilities besides palliative care work, leading to work overload and an inability to follow up with patients receiving palliative care.

> *“In terms of human resource, we need a focal person, an expert to guide us but we still don’t have the one for pediatrics! That affects us and that’s why we tend to rush quickly to the adult section to borrow someone, so it delays the process of administering timely care”**-KII 4, UCI***

Regarding human resources challenges, a few key informants noted that even where some healthcare workers skilled in palliative care existed, they were often engaged in general clinical care, which they found overwhelming. This left no time for palliative care activities such as patient follow-up, as one health worker explained.

> *“We have one palliative care nurse but instead of doing palliative care work, they gave her clinical work! Initially we had started palliative care work in oncology with a book to follow up patients but we gave up. We are overwhelmed. Others we forget to follow up because we are overwhelmed with work so the challenges are really many…”**-KII 3, MNRH***

#### Knowledge gap among health workers

Most key informants commented that, in as much as they could do PC assessment and offer care, they doubted if other health workers could do the same due to skill and knowledge gaps as one KII explained.

*“I feel there is knowledge gap among health workers! I told you I can do the palliative care assessment and but am not sure whether everyone else does it! We need to bridge that gap so that all health workers are able to consider palliative care as part of routine care. The other challenge would be staff! For you to be able to deliver good quality palliative care you need people who are dedicated to the service-palliative care alone, who will sit down with the patients and families at different phases of treatment.”-**KII 2, MNRH***

#### Lack of Palliative care prioritization

Most of the key informants reported to have, neither have PC units nor committees. These, together with the absence of a palliative care focal person limited their ability to follow up patients, after discharge.

> “*The cancer institute hasn’t generally gone far in terms of palliative care services because a facility like this is supposed to have a fully-fledged unit or department dedicated to handle the palliative care aspect but…”**-KII 4, UCI***

#### Limited access to services

Limited access to PC services due to frequent transfers of health workers and limited availability of morphine at the lower health facilities was another challenge mentioned to affect the continuity of PPC. Much as morphine is free of charge, many lower health facilities are not accredited to order it.

> *“Palliative care persons are stationed in the original district headquarters or in the original referral hospitals-those ones in HCIII or HCIV. You contact and they will tell you am no longer there we are not in existence. Morphine is cost-free but sometimes in those facilities, the medicines aren’t there. We will try and get 3 or 4 bottles of morphine put it on the bus”-**KII 4, UCI***

#### Stigmatization of Palliative Care

Participants mentioned the challenge of work and professional colleagues who stigmatize palliative care and demoralize those who practice it, thus hindering continuity as sometimes the PC providers are shifted from PC units to other departments like surgery.

> “*We are being fought when you want to do it [palliative care], our fellow medical workers say ‘why are you doing palliative care? That is useless!’ They even remove one from palliative care units and take them to surgery, medical because they think you are not performing! They don’t see it as something that is important, thus hindering continuity of the service”**-KII 1, MNRH***

#### Lack of child-friendly services

A few key informants noted the unavailability of child-friendly services. One of the palliative care providers at the RHHJ alluded to the fact that the facility has no child-friendly services thus children are not motivated to continue receiving care at that facility as they get frightened, thinking that they will get injections even when the facility only offers palliative care. The same space is used for both adults and children.

> *“We don’t have a specialized clinic for children! We only handle the children in the same clinic with the adults-there is no space so we end up sharing the same space –when you look around there are no child friendly services so when a child comes here it knows its injections yet we don’t inject here! But because they have been in those procedures, they come here very frightened and it’s really hard to relieve their fears, this hinders continuity of PC among the children”. –**KII 09 –RHHJ**.*

#### Navigation challenges

Key informants reported difficulty in navigation as a challenge that hinders the continuity of PC. Following discharge, patients get lost due to the inability to find the specific PC facility they are referred to for continuity of the service. This hinders the continuity of the service as one informant explained.

> *“People get lost-they can be given a referral from the cancer institute to hospice Jinja and the care taker fails to locate us. A patient may be discharged from the cancer institute and not sent to any specific palliative care facility they continue coming to UCI for review but when they aren’t aware of any palliative care centre where they can access care within that period thus they don’t continue with PC”-**KII 08-RHHJ.***

#### Poor service proximity resulting into high transport costs

Most of the key informants stated that, children stay far from the facility and its costly to transport them from home to the facility and this leads to non-continuity of PC.

*“We are also challenged in a way that it’s not easy for us to have children in one place – having children care in a particular place would be very good but because of the diverse distance it’s hard to collect one from Namayingo, two from Mayuge and bring them together so we are challenged! The fuel, you have to transport them and this gets costly for the organization”-**KII 08-RHHJ***

#### Communication challenges

Most of the key informants noted how difficult it is for children to continue PC when there is a communication barrier between that PC providers and the caregivers. Some of the caregivers do not have phones and yet for some, language barrier poses a great challenge. As patients don’t get to understand the health worker’s instructions in regards to continuity of palliative care.

> “*Most of them don’t even have phones and telephone numbers. There is one I really hustled, the telephone number in the file was for the neighbor who had even shifted to another place”-**KII 1, MNRH***

### Policy-related barriers influencing non-continuity of PC for children with cancer in Busoga sub-region

#### Inadequate funding/budgetary allocation for palliative care

Most of the key informants alluded to inadequate funds or government budgetary allocation towards PC as a key hindrance to the continuity of PPC. On some occasions, they noted to have used their own funds to follow-up patients in their homes or by phone calls.

> *“When the patients go home it’s really hard. There is no provision for following up or visiting patients at home. Sometimes, we really suffer as palliative care people; when there is a patient who needs home visiting, I go for a home visit, I pull money from my own pocket. Sometimes I follow them up using my money…”**-KII 1, MNRH***

**b) Facilitators**

### Health system-related facilitators influencing continuity of PC for children with cancer in Busoga sub-region

#### Care coordination and referral pathway-PC beyond and after discharge from the hospital

Most of participants narrated how they are able to coordinate care and link referrals, especially after discharge to ensure care continuity beyond the hospital; after the patient has been discharged home. Liaising with other palliative care providers, particularly hospices and using telephone follow-ups were mentioned as the common ways being utilized:

> *“The community based is of course more challenging but we try our level best through collaborations, phone call. When we get children from those particular areas where we have got collaborators, especially hospices to do for us the home visits, we coordinate. For example, for patients in Eastern Uganda, we coordinate Rays of Hope, Kitovu mobile in greater Masaka… We facilitate the provision of painkillers particularly morphine and we get a lot of help from hospice Uganda. Sometimes we may have to put the morphine on private means or a taxi and send it to the community when the patient is stuck there without morphine” **-KII 2, MNRH***

## DISCUSSION

### Prevalence of Non-Continuity of Palliative Care for Children with Cancer in the Busoga Sub-Region

This study found that 74 out of 77 (96.1%) children with cancer in the Busoga sub-region experienced non-continuity of palliative care (PC). This high prevalence indicates that most children from this region miss out on the benefits of PC following discharge from in-patient care. As a result, many of these children continue to suffer from pain, psychological, and spiritual challenges, and those at the end of life may not experience a quality death, as most are likely to die in pain. The prevalence of non-continuity was almost evenly distributed between the two tertiary facilities: 37 (97.4%) at MNRH-PHOU and 37 (94.9%) at UCI. This similarity may be due to the lack of designated PPC teams at both facilities, with PPC experts working independently and patient follow-up relying on individual initiative. The outdated staffing structures of the two facilities, which did not accommodate palliative care positions, likely limit the number of children who can be supported with morphine for continued PC. The prevalence in this study is significantly higher than the 79% reported by Rehner et al. [14] in Mecklenburg-Western Pomerania and aligns with another Ugandan study indicating that only 10% of those needing PC generally receive it [15]. This high prevalence is partly due to the lack of integration of pediatric palliative care into Uganda’s healthcare system, despite its introduction over 20 years ago. Initially focused on adults with advanced HIV disease, PC services have declined with the scale-up of ART, which transformed HIV into a chronic disease. Currently, pediatric palliative care (PPC) is available mainly at a few tertiary facilities with limited linkage to community-based services, hindering continuity post-discharge when patients return to distant homes.

### Barriers to Continuity of PPC in the Busoga Sub-Region

#### Individual Level

Caregivers’ reluctance to continue PC was often attributed to cultural beliefs such as attributing cancer to witchcraft or supernatural punishment for wrongdoing. Such beliefs hinder the continuity of PC, leading to unmanaged pain and other challenges. This finding is consistent with Hawley [16], which reported reluctance to accept palliative care referrals due to cultural beliefs in Denmark. Similarly, a study in Ethiopia found that patients’ preference for conventional medicine was a barrier to PC services [17].

#### Relationship Level

The study revealed that a cancer diagnosis often leads to social challenges such as parental separation or divorce, which disrupts the necessary support system. This family breakdown can leave the child vulnerable to non-continuity of PC, as single parents may struggle with transport and emotional support. Mader et al. [18] found that parents under 45 years who were unemployed prior to the child’s cancer diagnosis had a higher risk of separation or divorce. Haimi and Lerner [19] also highlighted that divorce can exacerbate children’s mental health issues and increase non-continuity of PC. Given that cancer diagnosis is associated with most of these mental challenges, parent separation without full family support, interruptions in care-seeking become inevitable due to issues like difficulty in raising finances.

#### Health System Level

The study identified several healthcare system barriers, including limited human resources, inadequate training, and insufficient funding for palliative care. The shortage of skilled personnel in PPC and the lack of training and expertise could be attributed to inadequate funding and low interest among medical personnel in pediatric palliative care due to its emotional challenges. Limited human resources affect service continuity, as there is inadequate follow-up and a risk of missing referral information. Similar challenges were noted by Mosoiu et al. [20] in Romania and Abate et al. [21] in Ethiopia, including turnover, shortage of healthcare workers, and lack of government support. Insufficient training of health professionals in LMICs has also been reported as a major obstacle [22]. Increasing the number of trained PPC providers could alleviate workload issues and enhance patient follow-up.

#### Societal Level

Structural issues such as the absence of a national palliative care policy lead to inconsistent service delivery, inadequate resource allocation, and limited advocacy and research. A national palliative care policy is crucial for guiding the management of PC, ensuring strategic prioritization, and mobilizing resources. The WHO’s 2016 public health approach to palliative care emphasized the need for such policies [23]. Policies like Kenya’s PC policy, launched in 2021, provide better guidance for resource allocation and service delivery [24].

### Facilitators to Continuity of PPC in the Busoga Sub-Region

#### Individual Level

Caregivers’ understanding of palliative care significantly facilitated its continuity. All caregivers of children who continued to receive PC had some level of understanding of PC, while those who did not continue had no information about it. This finding aligns with Hudson et al.,2019 which emphasized that informed caregivers contribute to effective care [27]. The WHO guide for effective PC programs supports the identification and training of community and family caregivers in low-resource settings [26].

#### Health System Level

Training and prior knowledge of PC among healthcare providers were significant facilitators. Key informants with training in palliative care were better equipped to provide continuous care, including pain management and psychosocial support. This finding is consistent with Cai et al.,2022 in China, which linked limited training to deficiencies in PC competencies [26]. Coordinated care, particularly post-discharge, involving referrals and follow-ups, was crucial for continuity. This is supported by Hudson et al., 2019 and Fyeza et al. which highlighted the importance of education and regular follow-ups for PC continuity [27].

#### Study Strengths

study represents the first investigation into non-continuity of pediatric palliative care in Uganda and is among the few studies on pediatric palliative care in East Africa.

#### Study Limitations

The high mortality rate limited the population from which a sample could be drawn, making it impossible to perform a multivariate statistical analysis. Additionally, recall bias may have affected the accuracy of information, as participants had to remember what they were told upon discharge. Selection bias was also a concern, as those who had died were excluded from the study. Furthermore, some confounders could not be measured or controlled due to the retrospective nature of the study.

## Conclusion

The study highlights an unacceptably high prevalence of non-continuity of palliative care for children with cancer. This suggests that the integration of palliative care into pediatric cancer care in Uganda is still insufficient, revealing a systemic gap between facility-based and community-based palliative care providers. Health system-related factors predominantly influence non-continuity. Without a comprehensive understanding of pediatric palliative care by health workers and the establishment of an appropriate policy environment, non-continuity of palliative care for children with cancer is likely to persist.

There is urgent need to develop a national palliative care policy, strategy, and guidelines to standardize and improve palliative care services. Health facilities should ensure that palliative care providers issue a discharge plan that includes steps for continuity of care. Empower and support patients to facilitate their own continuity of palliative care by mapping local palliative care providers who can continue care post-discharge. Integrate palliative care into pediatric oncology by establishing dedicated palliative care teams in cancer treatment facilities.

## Data Availability

All data underlying this study is fully available as a complete thesis at Makerere University physical and online libraries

## Acknowledgments

Appreciation to Mulago National Referral Hospital, UCI and Rays of Hope-Hospice, Jinja (RHHJ), for allowing us access to the databases and letting their staff participate in the study.

Much gratitude to all the participants, for consenting to participate in this study. Many thanks to Dr. Eve Namisango for the support during qualitative data analysis

## Notes

### Competing Interest Statement

The authors have declared no competing interest.

### Funding Statement

The study was sponsored by the Palliative Care Association of Uganda (PCAU) and the African Palliative Care Association (APCA). The Palliative Care Association of Uganda (PCAU) was established in 1999 and registered as a Non-Governmental Organization (NGO) in 2003. PCAU was established to coordinate civil society efforts in supporting the government to integrate palliative care into the national health care system at all levels. PCAU was composed of 26 organizations and over 1300 individual members that included health care professionals, caregivers, and community members across the country. The association’s secretariat operates from owned premises in Kitende on Entebbe Road and runs 10 regional committees (branches) in the country. One of the core objectives of PCAU is to establish a hub of research and information on palliative care in Uganda. This study will contribute to the body of research sought for improving access and continuity to palliative care in Uganda. PCAU facilitated this study with 1300 USD to support data collection and analysis. APCA was established after a meeting in Cape Town in 2002 of 28 palliative care trainers from across Africa. The group produced the Cape Town Declaration, which holds palliative care and pain and symptom control as a human right for every adult and child with life-limiting illnesses. APCA supported the study with 800 USD that complimentary to the funding from PCAU. This was also used to facilitated data collection and analysis.

### Author Declarations

Makerere School of Medicine Research and Ethics Committee (SOMREC)

